# Antiviral metabolite 3’-Deoxy-3’,4’-didehydro-cytidine is detectable in serum and identifies acute viral infections including COVID-19

**DOI:** 10.1101/2021.07.23.21260740

**Authors:** Ravi Mehta, Elena Chekmeneva, Heather Jackson, Caroline Sands, Ewurabena Mills, Dominique Arancon, Ho Kwong Li, Paul Arkell, Timothy M. Rawson, Robert Hammond, Maisarah Amran, Anna Haber, Graham Cooke, Mahdad Noursadeghi, Myrsini Kaforou, Matthew Lewis, Zoltan Takats, Shiranee Sriskandan

**Affiliations:** Department of Infectious Disease, Imperial College London, UK; The National Phenome Centre, Imperial College London, UK; Division of Systems Medicine, Department of Metabolism, Digestion and Reproduction, Imperial College London, UK; Imperial College Healthcare NHS Trust, UK; MRC Centre for Molecular Bacteriology & Infection, Imperial College London, UK; Division of Infection & Immunity, University College London, UK; NIHR Health Protection Research Unit in Healthcare-associated Infection & Antimicrobial Resistance, UK

## Abstract

There is a critical need for improved infectious disease diagnostics to enable rapid case identification in a viral pandemic and support targeted antimicrobial prescribing. Here we use high-resolution liquid chromatography coupled with mass spectrometry to compare the admission serum metabolome of patients attending hospital with a range of viral infections, including SARS-CoV-2, to those with bacterial infections, non-infected inflammatory conditions and healthy controls. We demonstrate for the first time that 3’-Deoxy-3’,4’-didehydro-cytidine (ddhC), a free base of the only known human antiviral small molecule ddhC-triphosphate (ddhCTP), is detectable in serum. ddhC acts as an accurate biomarker for viral infections, generating an area under the receiver operating characteristic curve of 0.954 (95% confidence interval 0.923-0.986) when comparing viral to non-viral cases. Gene expression of viperin, the enzyme responsible for ddhCTP synthesis, is highly correlated with ddhC, providing a biological mechanism for its increase during viral infection. These findings underline a key future diagnostic role of ddhC in the context of pandemic preparedness and antimicrobial stewardship.

## Introduction

Early differentiation of acute infectious aetiologies is now a priority in diagnostic innovation. Conventional methods relying on pathogen identification through culture, polymerase chain reaction or antigen detection are time-consuming and/or insensitive, leading to diagnostic delays that result in inappropriate antimicrobial prescription and infection transmission.^1-3^ There is therefore renewed interest in novel biomarkers of infection classes that can better guide therapeutic and infection control decisions in real-time.

Metabolomics technologies for large-scale characterisation of low-molecular-weight metabolites have the potential to aid discovery of novel biomarkers of infectious diseases. Liquid chromatography coupled with mass spectrometry (LC-MS) and nuclear magnetic resonance (NMR) spectroscopy stand out among the most commonly employed techniques in the field. The use of mass spectrometry has already revolutionised modern microbiology by enabling rapid detection of bacterial species from cultured colonies.^4^

Despite its growing impact on biomedical research, metabolic profiling of biofluids has produced candidate biomarkers in only a small number of infectious states. One study identified a two-metabolite serum signature differentiating infected from non-infected patients within a systemic inflammatory response syndrome cohort.^5^ Metabolomic interrogation of cerebrospinal fluid from patients with meningitis was able to differentiate between *M. tuberculosis* and other infectious causes.^6^ Wang *et al*. examined the lipidome of 40 patients in a paediatric cohort prior to the COVID-19 pandemic and identified a 3-lipid signature that discriminated bacterial from viral infection, although wider metabolomic changes were not reported.^7^ A number of more recent studies report metabolic differences between patients with and without SARS-CoV-2 infection,^8-10^ but comparator groups did not include bacterial infections.

We investigated the serum metabolome of adult patients presenting to two UK emergency departments with a range of infections, including SARS-CoV-2, to derive and cross-validate novel biomarkers for viral and bacterial infections. We used point-of-admission samples to replicate the timepoint where a discovered biomarker would be used clinically. To ensure diagnostic certainty, we adopted a case-control approach with laboratory-proven infections. Our sampling included unwell, non-infected cases to ensure that any biomarkers identified accounted for cases of inflammatory conditions unrelated to infection.^11^

## Methods

### Study population

Serum samples from adult patients presenting to the emergency department were obtained from two parallel studies at a major North-West London Teaching Hospital Trust, the Bioresource in Adult Infectious Diseases (BioAID)^12^ and Microbial Products in Infection, from August 2014 – December 2020.

Patients were recruited to BioAID if they had a suspected clinical infection syndrome of sufficient severity, as assessed by a clinician, to warrant blood culture testing. Blood samples were obtained at the point of admission, alongside microbial isolates identified during the inpatient stay, in conjunction with demographic and clinical data. Ethical approval was obtained to take deferred consent from patients (or next of kin/nominated consultee) to retain blood samples, including serum and RNA specimens, as well as clinical data (Research Ethics Committee [REC] references 14/SC/0008 and 19/SC/0116).^12^

Patients were retrospectively identified as part of the Microbial Products in Infection protocol where a pathogen of interest – in this study, SARS-CoV-2 – was identified to the research team by the routine diagnostic laboratory. Serum samples obtained at the point of admission were linked to anonymised demographic data including age, sex, timing of sample in relation to illness onset, survival/death, antibiotic use, ICU admission/duration of stay, and blood test results. (REC reference 06/Q0406/20).

For both studies, all serum samples were taken in the same manner at the point of patient admission to hospital, prior to any intervention, as part of usual clinical care. Following routine diagnostic testing, surplus volumes were retrieved from the Clinical Chemistry laboratory where they were stored at 4°C, and transferred to a −80°C freezer within five days of sample acquisition. During the peaks of the UK COVID-19 pandemic (March-April and September-December 2020), owing to unprecedented pressures placed on laboratory staffing, some serum samples from COVID-19 patients (n=80) were transferred to a −80°C freezer between six and 30 days from sample acquisition and were classified into a separate sub-group – these were not used in the primary analysis to avoid confounding bias. Control serum samples from consenting healthy donors were from an approved subcollection of the Imperial College Healthcare NHS Trust Biomedical Research Council (ICHT BRC) Tissue Bank (approval R12023).

### Patient selection

Serum samples from patients in one of the following six categories were used in this study: Gram-positive bacteraemia, Gram-negative bacteraemia, PCR-confirmed viral infection prior to detection of SARS-CoV-2 in the UK (January 2020), PCR-confirmed COVID-19, non-infected patients, and healthy controls.

N=24 samples in each of two comparator groups were required to achieve a power of >90% to identify an AUC of at least 0.8, at a significance level of 0.01. Thus to enable all comparisons, accounting for potential sample exclusion (e.g. assay failure, poor data quality), we used n=30 samples in each clinical group apart from COVID-19, where we included all available samples (n=112) to facilitate exploration of severity differences in this cohort in future work. Infection categories were assigned using electronic diagnostic pathology data pertaining to admission only and admission case notes were cross-checked for diagnoses by a clinician. Non-infected patients were identified from the database where there was no positive microbial diagnostic test and no infection-related ICD-10 diagnostic code from BioAID admission. Sera from n=13 healthy controls were available from the ICHT BRC Tissue Bank.

To facilitate multi-omic comparison, serum samples from BioAID patients were prioritised if whole blood RNA-Sequencing had already been undertaken as part of an earlier study, where samples had been selected from the BioAID database using a random number generator.^13^ Sera from additional BioAID patients were selected randomly from within individual infection groups using a random number generator in Excel.

Bacteraemic patients were excluded if the isolated bacterium was deemed a contaminant, or if the blood culture was taken >24 hours prior to/post the admission serum sample. COVID-19 patients were excluded if their positive PCR test was taken >10 days prior to admission, or >2 days post admission, to avoid non-COVID-19 related admissions and hospital-acquired COVID-19, respectively. Co-infections across different infection classes were also excluded.

### Metabolic profiling

Serum samples from 245 patients were analysed using ultra-performance LC-MS following previously described analytical and quality control (QC) procedures.^14^ A suite of chromatographic separations was used, each coupled with high resolution time of flight mass spectrometry, to maximise coverage of a broad range of metabolite and lipid classes. Hydrophilic interaction liquid chromatography (HILIC) was used for the separation of hydrophilic analytes (e.g. polar and charged metabolites), while reversed-phase chromatography (RPC) was used for the separation of lipophilic analytes to profile complex and neutral lipid species.^15^ Each RPC LC-MS assay was conducted in both negative and positive ionisation modes, producing lipid RPC- and lipid RPC+ datasets. The HILIC LC-MS assay was conducted in the positive ionisation mode only, producing the HILIC+ dataset.

For each assay, samples were analysed in a randomised order demonstrating no correlation or other relationship with study design variables, precluding any confounding effect of analysis order. To facilitate quality assessment and pre-processing, a pooled QC sample was prepared by combining equal parts of each study sample and analysed periodically among study sample analyses. In addition, for assessment of analyte response,^16^ a series of QC sample dilutions was created (10 × 100%, 5 × 80%, 3 × 60%, 3 × 40%, 5 × 20%, 10 × 10%, 10 × 1%) and analysed at the start and end of each set of sample analyses.

Serum samples were prepared as previously described.^15^ In brief, 50⍰μL aliquots were taken from each sample and the pooled QC and diluted 1:1 v/v with ultrapure water. Protein was removed by addition of organic solvent (diluted sample/isopropanol in 1:4 v/v ratio for lipid RPC profiling and diluted sample/acetonitrile in 1:3 v/v ratio for HILIC+ profiling). Mixtures of method-specific authentic chemical standards were added at the dilution step (HILIC) or the protein precipitation step (lipid RPC) in order to monitor data quality during acquisition. Sample analyses were performed on ACQUITY UPLC instruments (Waters Corp., Milford, MA, USA) coupled to Xevo G2-S Q-TOF mass spectrometers (Waters Corp., Manchester, UK) via a Z-spray electrospray ionisation (ESI) source operating in either positive or negative ion mode.

Raw data were converted to the mzML open-source format and signals below an absolute intensity threshold of 100 counts were removed using the MSConvert tool in ProteoWizard ^17^ before data extraction using XCMS,^18^ outputting a matrix of measurements (peak integrals) organised row-wise into samples and column-wise into LC-MS “features”, each of which is described by its mass:charge (*m/z*) value and chromatographic retention time. All datasets were further processed using the nPYc-Toolbox^19^ for elimination of potential run-order effects and filtering of features not meeting previously established QC criteria. Only features measured with high analytical quality (RSD in pooled QC<30%, pooled QC dilution series Pearson correlation to dilution factor>0.7, RSD in study samples>1.1* RSD in pooled QC) were retained and put forward for further statistical analysis. After feature filtering, datasets contained the following number of variables; lipid RPC-: 521; lipid RPC+: 2257; HILIC+: 1572.

### Metabolite annotation

The molecular formula of a feature of interest was determined by elemental composition analysis of the high resolution time of flight data obtained in the metabolic profiles of the study samples. Matches for the observed *m/z* values were sought in chemical and spectral databases (Human Metabolome Database,^20^ METLIN,^21^ NIST17,^22^ Mass Bank of North America [http://massbank.us/]). A putative identity was assigned based on the comparison of the observed accurate mass and collision-induced dissociation (CID) tandem mass spectrometry (MS/MS) fragmentation data to that available in published literature.^23^ The HILIC chromatographic method used for CID-MS/MS analysis was the same as that used in the profiling method with the same MS source conditions.^14^ CID-MS/MS target selection was performed using unit mass selection via quadrupole with a collision energy voltage ramp of 10-45V. The metabolite identity was validated by matching the accurate mass, isotope distribution, fragmentation pattern, and retention time obtained from the feature observed in study samples to those obtained by analysis of a chemical standard (acquired from Berry & Associates). The full procedure for the identified biomarker is described in detail in the Supplementary.

### Statistical analysis

Data analysis was performed using R.^24^ Power calculations were performed using the pROC package.^25^ Unit-variance scaled principal component analysis (PCA) and eigencor plots were performed on the primary analysis cohort to identify the major sources of variation in the dataset, using the PCAtools package.^26^ For PCA, features where >98.5% of samples returned an intensity of zero were excluded (n=3/1572 in HILIC+ dataset, nil in both lipidomics datasets).

We compared all viral cases (COVID-19 and pre-COVID-19) versus others, all bacterial cases (Gram-positive and Gram-negative bacteraemia) versus others, and all viral versus all bacterial cases. In each comparison, we assessed the fold-change between the infection groups’ median intensities for each feature. P-values were generated using the two-sided Wilcoxon test and were adjusted using the Benjamini-Hochberg procedure.^27^ Volcano plots were generated comparing median log_2_fold-change and -log_10_ p-values.

In order to cross-validate findings, we used the variable selection method, forward selection-partial least squares (FS-PLS).^28^ FS-PLS has been described in detail elsewhere.^7,29^ Briefly, it is a forward-selection method that selects variables most strongly associated with the groups of interest. It can be used to select a multi-feature signature composed of non-correlated variables, but in this study the ‘max’ parameter was set to one to evaluate the performance with only one feature. Feature intensities were log_2_ transformed. A p-value threshold of 0.01 was used, which determined the selection of a variable or termination. 100 runs of FS-PLS were applied to the dataset for every comparison, each time with a different training:test split at a ratio of 70:30. In each FS-PLS run, the feature identified on the training set was tested on the test set, and its performance was assessed using the AUC generated. For the feature that was selected in the most FS-PLS runs out of 100, the median and interquartile range (IQR) of the respective test AUCs were generated.

To assess the diagnostic utility of features of interest and compare it to the traditional biomarkers C-reactive protein (CRP), white cell count and lymphocyte count (procalcitonin levels were not routinely available), AUCs were generated for the entire primary analysis cohort using the pROC package.^25^ The Youden’s J statistic was used to determine thresholds for sensitivity and specificity.^30^

### Multi-omic comparison

We examined the interaction between whole blood gene expression and the feature of interest identified in this study. Gene expression data were obtained from RNA-Sequencing (RNA-Seq) of BioAID patient RNA samples, performed prior to this study in two cohorts. Full details for the first patient cohort (recruited pre-COVID-19 pandemic) have been described previously (Li. *et al*, supplementary p. 3).^13^ For the second cohort (recruited during the COVID-19 pandemic), whole blood was collected in the same way as the first cohort. Material was quantified using RiboGreen (Invitrogen) on the FLUOstar OPTIMA plate reader (BMG Labtech) and the size profile and integrity analysed on the 2200 TapeStation (Agilent, RNA ScreenTape). Input material was normalised and strand specific library preparation was completed using NEBNext® Ultra™ II mRNA kit (NEB) and NEB rRNA/globin depletion probes following manufacturer’s instructions. Libraries were on a Tetrad (Bio-Rad) using in-house unique dual indexing primers (based on Lamble et al).^31^ Individual libraries were normalised using Qubit and pooled together. The pooled library was diluted to ∼10 nM for storage and denatured and further diluted prior to loading on the sequencer. Paired end sequencing was performed The Wellcome Centre for Human Genetics in Oxford UK using a Novaseq6000 platform at 150 paired end configuration, generating a raw read count of 30 million reads per sample. The RNA-Seq analysis pipeline consisted of quality control using FastQC,^32^ MultiQC^33^ and annotations modified with BEDTools,^34^ alignment and read counting using STAR,^35^ SAMtools,^36^ FeatureCounts^37^ and version 89 ensembl GCh38 genome and annotation.^38^

Genes completely missing in either of the RNA-Seq cohorts were removed, in addition to ribosomal genes. The two RNA-Seq cohorts were merged and the batch effects between the two cohorts, in addition to the plate effects within the first cohort, were removed by combat_seq.^39^ The raw counts were normalised using DESeq2.^40^ In patients for whom both metabolic and transcriptomic data were available, we assessed the correlation between log_2_-transformed feature intensities of a metabolite of interest and log_2_-transformed expression of associated genes using Pearson correlation coefficients. We restricted further analysis to the five genes most highly correlated to the metabolite of interest.

## Results

### Samples

Serum from 232 patients and 13 healthy controls underwent metabolomic profiling. 173 patients were selected from the BioAID study (30 Gram-positive bacteraemia, 30 Gram-negative bacteraemia, 30 pre-COVID-19 viral, 53 COVID-19, 30 non-infected unwell control), 59 from Microbial Products in Infection (all COVID-19), 13 were healthy controls. Owing to insufficient sample volume or data quality, four samples were excluded from the primary analysis in both lipidomics assays, and five from the HILIC+ assay. The sub-group of 80 COVID-19 samples not transferred to a −80°C within five days of collection was also excluded from the primary analysis (Figure 1). In all assays, PCA did not show clustering of samples by age or sex and eigencor plots did not show correlation above 0.4 (Supplementary Figure 1). The final number of samples included in the primary analysis ranged from 161-163, depending on the assay, shown alongside patient demographics in Supplementary Table 1. Confirmed infection pathogens are shown in Supplementary Table 2.

**Figure 1.**
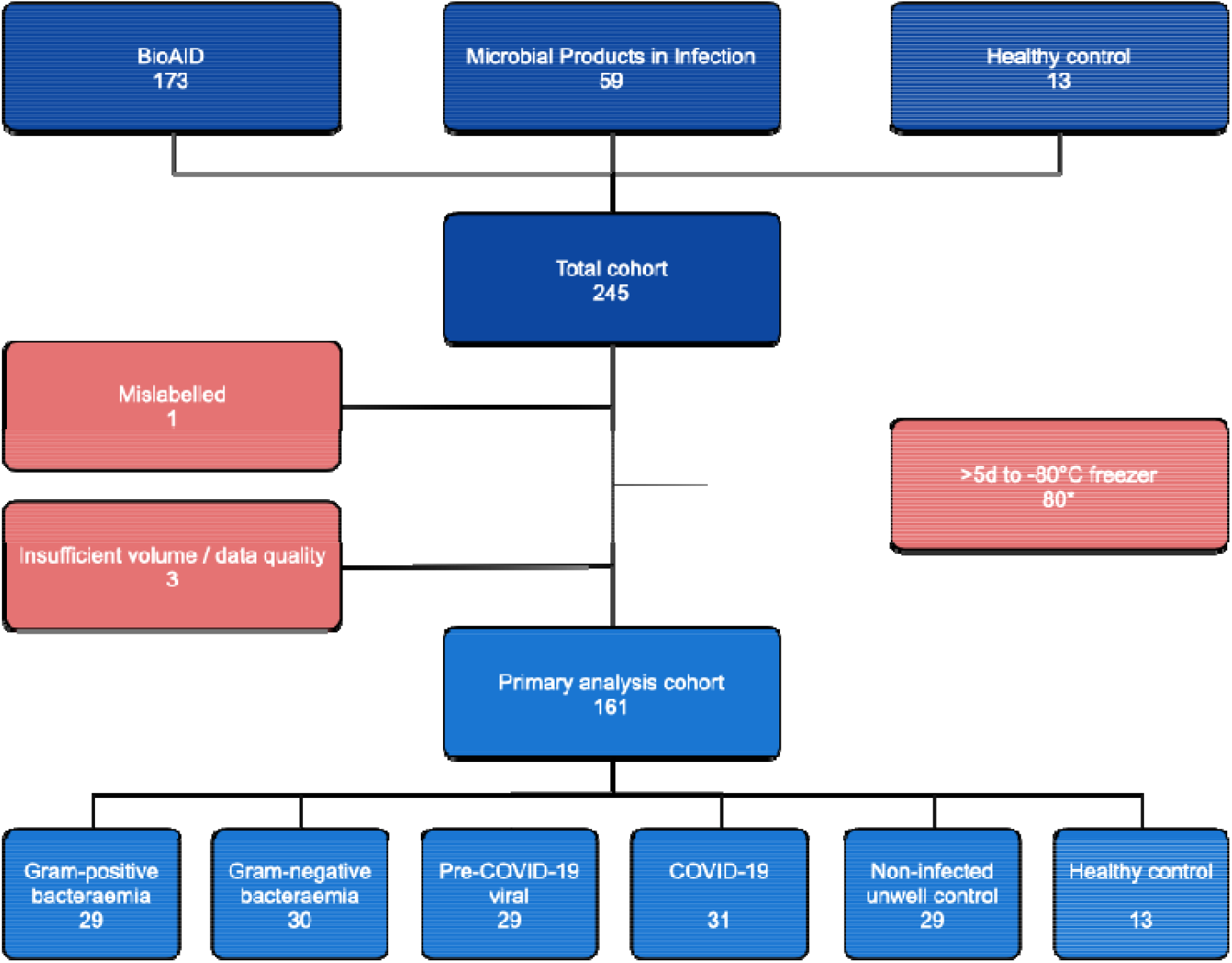
Flowchart of sample selection and exclusion for the primary analysis cohort, data shown for the hydrophilic interaction chromatography (HILIC+) assay. 80 COVID-19 samples were excluded as they were transferred to a −80°C freezer >5 days after collection (*two samples excluded within this group also had insufficient volume / data quality).

### Best performing discriminator for viral infections

Analysis of the HILIC+ dataset identified significantly differentially abundant (SDA) features with a median absolute log_2_ fold-change of >4 and p-value <0.01 when comparing viral cases (pre-COVID-19 viral and COVID-19) versus all other groups, and viral versus bacterial cases (Gram-positive and Gram-negative bacteraemia) (Figure 2). Using these empirical thresholds, no SDA features were identified in bacterial cases versus all other groups in the HILIC+ dataset, as well as in all comparisons in both lipidomics datasets (Supplementary Figure 2). The top SDA discriminator was the feature 248.0647 *m/z* at 1.96 minutes, which showed a 36-fold change in the median intensity in viral cases compared to all other groups (adjusted p-value <1×10^18^). The marker was identified as 3’-Deoxy-3’,4’-didehydro-cytidine (ddhC; Supplementary page 13), a free base of the antiviral ribonucleotide ddhC-triphosphate (ddhCTP) previously reported in the literature. ^41^

**Figure 2.**
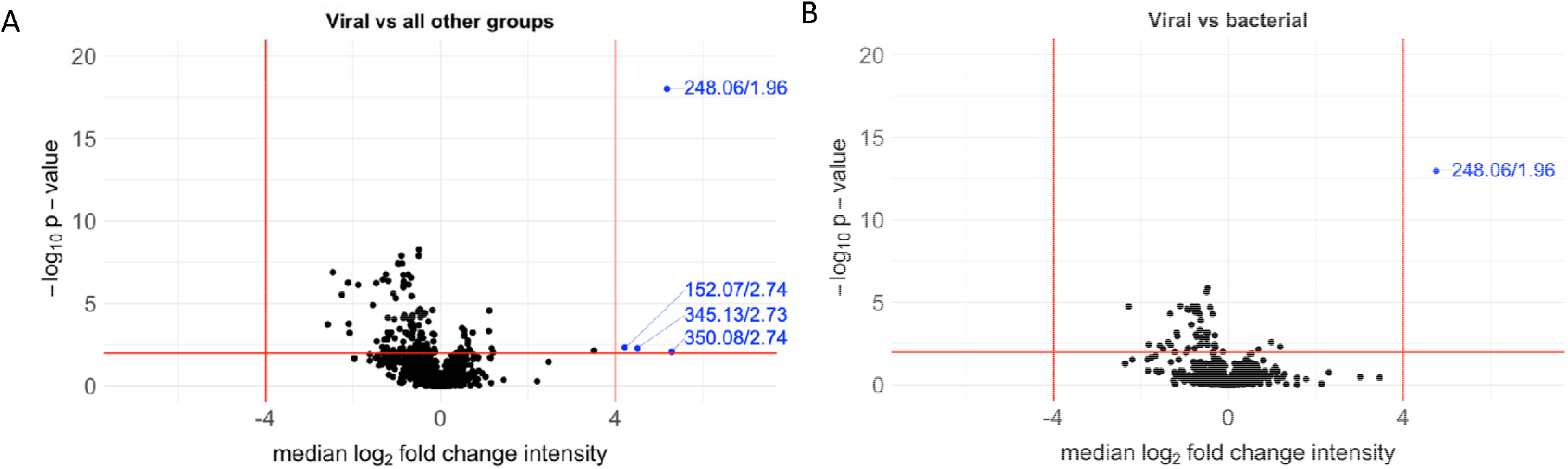
Volcano plot showing median log_2_ fold change in intensity of each feature versus -log_10_ p-value in the HILIC+ dataset (n=161) when comparing: **A**. viral cases (pre-COVID-19 viral and COVID-19) versus all other groups and **B**. viral versus bacterial (Gram-positive and Gram-negative bacteraemia) cases. Empirical threshold lines in red represent a fold-change of 16 [log_2_(foldchange) of 4] and p-value of 0.01 [-log_10_(p-value) of 2]. Candidate biomarkers are shown in blue by mass:charge ratio/retention time, with 248.06/1.96 (ddhC) performing best.

Using all samples from the primary analysis cohort, ddhC returned an AUC of 0.954 (95% CI 0.923-0.986; sensitivity 88.1%, specificity 91.7%) in discriminating viral infections from all other groups, and 0.944 (95% CI 0.905-0.983; sensitivity 89.8%, specificity 86.7%) in discriminating viral from bacterial infections (Figure 3). When we included the sub-group of samples that spent more than five days outside a −80°C freezer, similar results were achieved with AUCs of 0.966 and 0.959 respectively (Supplementary Figure 3).

**Figure 3.**
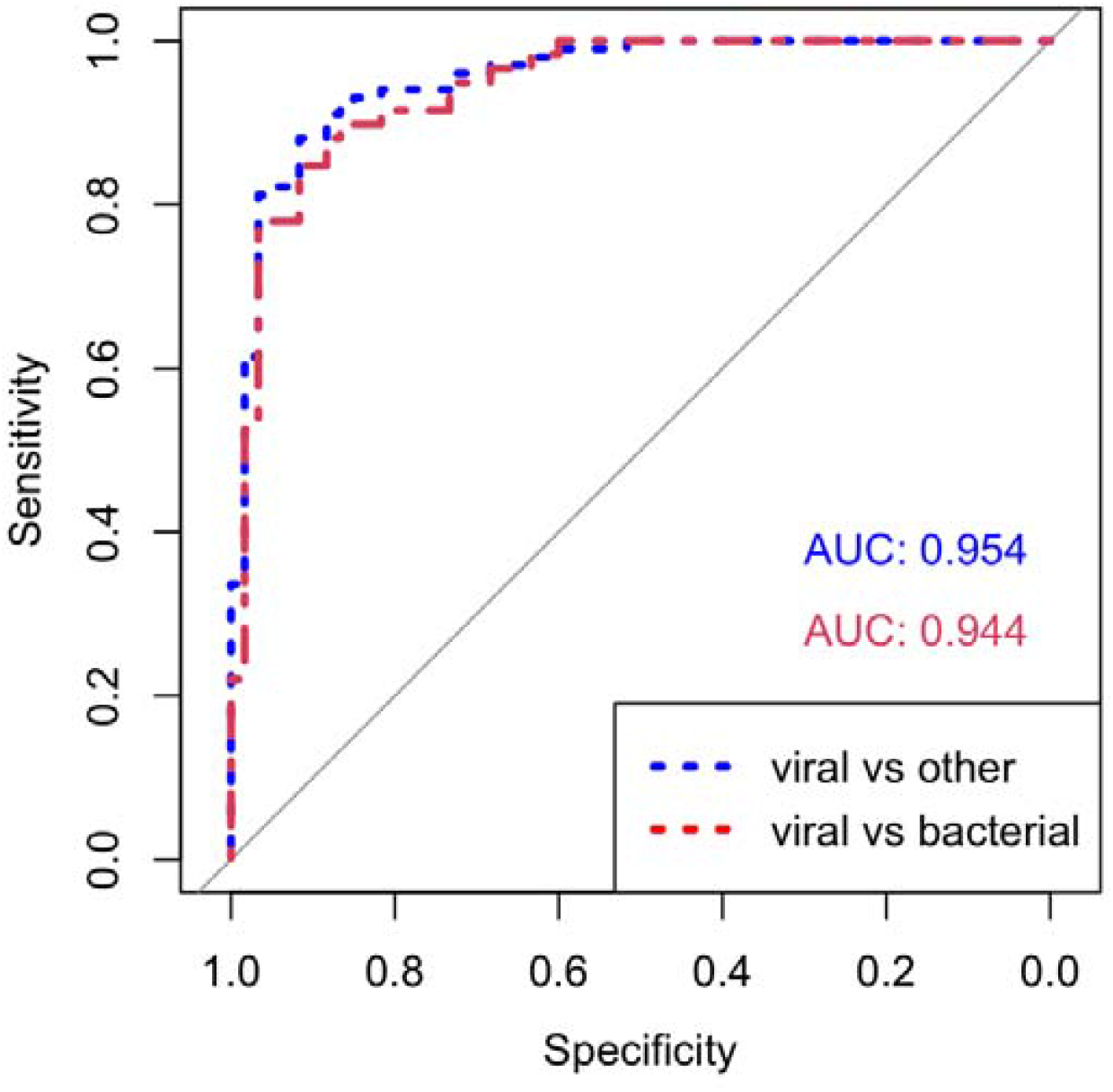
Area under the receiver operating characteristic curves (AUCs) for ddhC distinguishing viral versus other and viral versus bacterial groups. Using all samples in the primary analysis (n=161): **Blue**. AUC of 0.954 (95% CI 0.923-0.986) for ddhC differentiating viral infections from all other groups. **Red**. AUC of 0.944 (95% CI 0.905-0.983) for ddhC differentiating viral from bacterial infection (controls omitted).

Within the primary analysis cohort, ddhC demonstrated a higher relative intensity among patients with viral infections compared to other groups (Figure 4). Similar results were achieved when including samples that spent more than five days outside a −80°C freezer (Supplementary Figure 4). Within the viral group, there was no significant (p-value <0.01) difference in the median intensity of ddhC between age or sex subgroups (Supplementary Figure 5).

**Figure 4.**
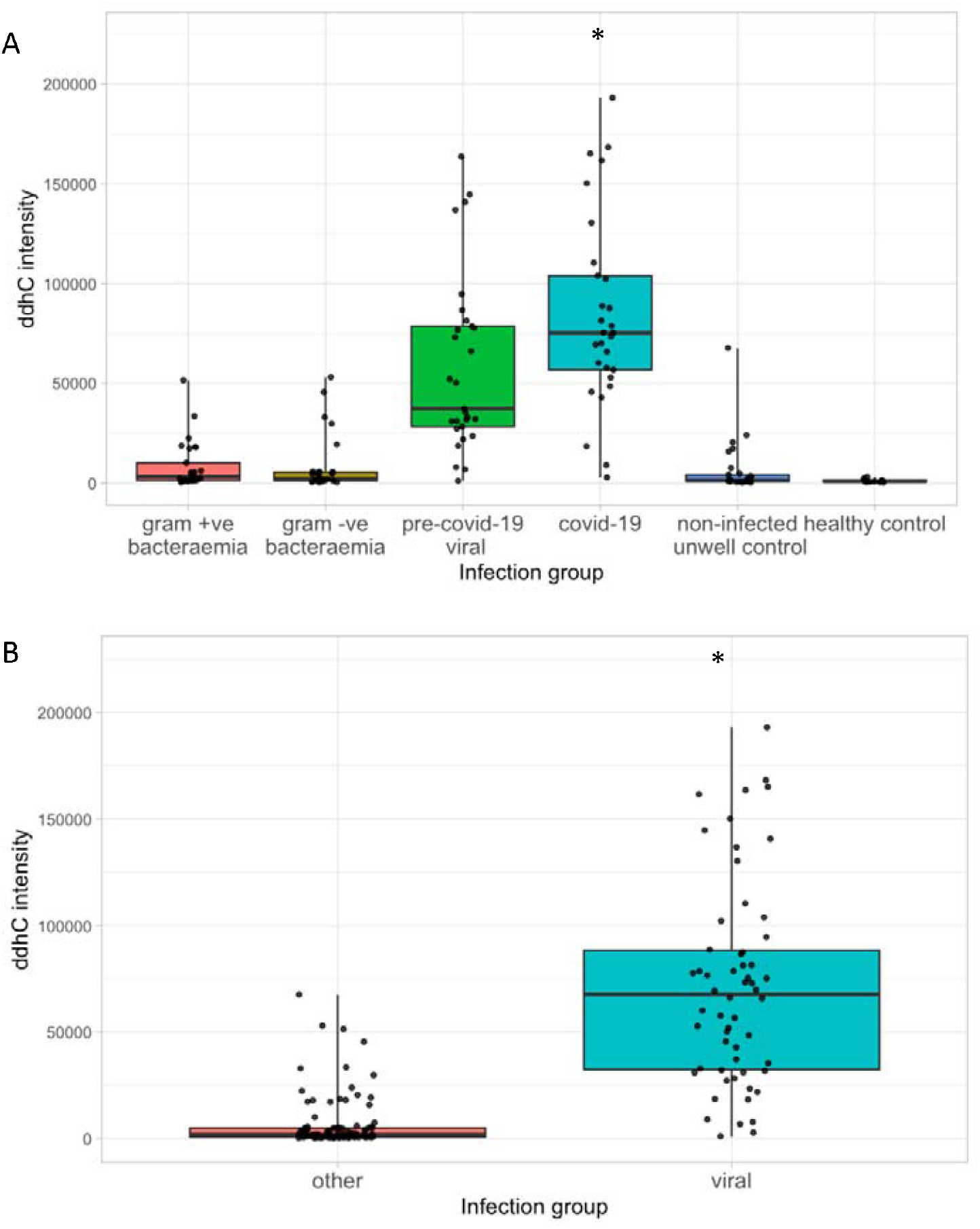
Relative ddhC intensity data in different patient groups. Points represent individual patients. Boxes represent interquartile ranges with medians. **A**. All comparator groups. **B**. Pre-COVID-19 viral and COVID-19 grouped together into one ‘viral’ group vs all other groups. *2 samples in the COVID-19 group had a relative intensity of >700000, not shown.

### Cross-validation using single-feature FS-PLS

To assess and cross-validate the discriminatory performance of single markers distinguishing infection groups in the data, we utilised the FS-PLS method. For each comparison (viral versus other, viral versus bacterial, bacterial versus other) we present the discriminating feature that was selected most frequently out of 100 different training:test FS-PLS runs and the median and IQR of the test AUCs generated (Supplementary Table 3). When comparing viral versus all other groups, ddhC (HILIC+, 248.06/1.96) was selected in all 100 FS-PLS runs, and generated a median (IQR) test AUC of 0.957 (0.943-0.970). When comparing viral versus bacterial groups, ddhC (HILIC+, 248.06/1.96) was selected in 99/100 FS-PLS runs, generating a median (IQR) test AUC of 0.951 (0.926-0.971).

### Comparison of ddhC as a biomarker to white cell count, lymphocyte count & CRP

We compared the ability of ddhC to differentiate viral infection from other groups to patients’ white cell count, lymphocyte count and CRP, which were taken as part of routine admission clinical laboratory tests. We used the primary analysis HILIC+ cohort (n=161) and excluded healthy controls (n=13), for whom there was no routine laboratory test data (n=148 in total). All patients had a white cell count and lymphocyte count recorded, and 122/148 patients had a CRP. Routine admission clinical laboratory tests performed poorly compared to ddhC (Figure 5).

**Figure 5.**
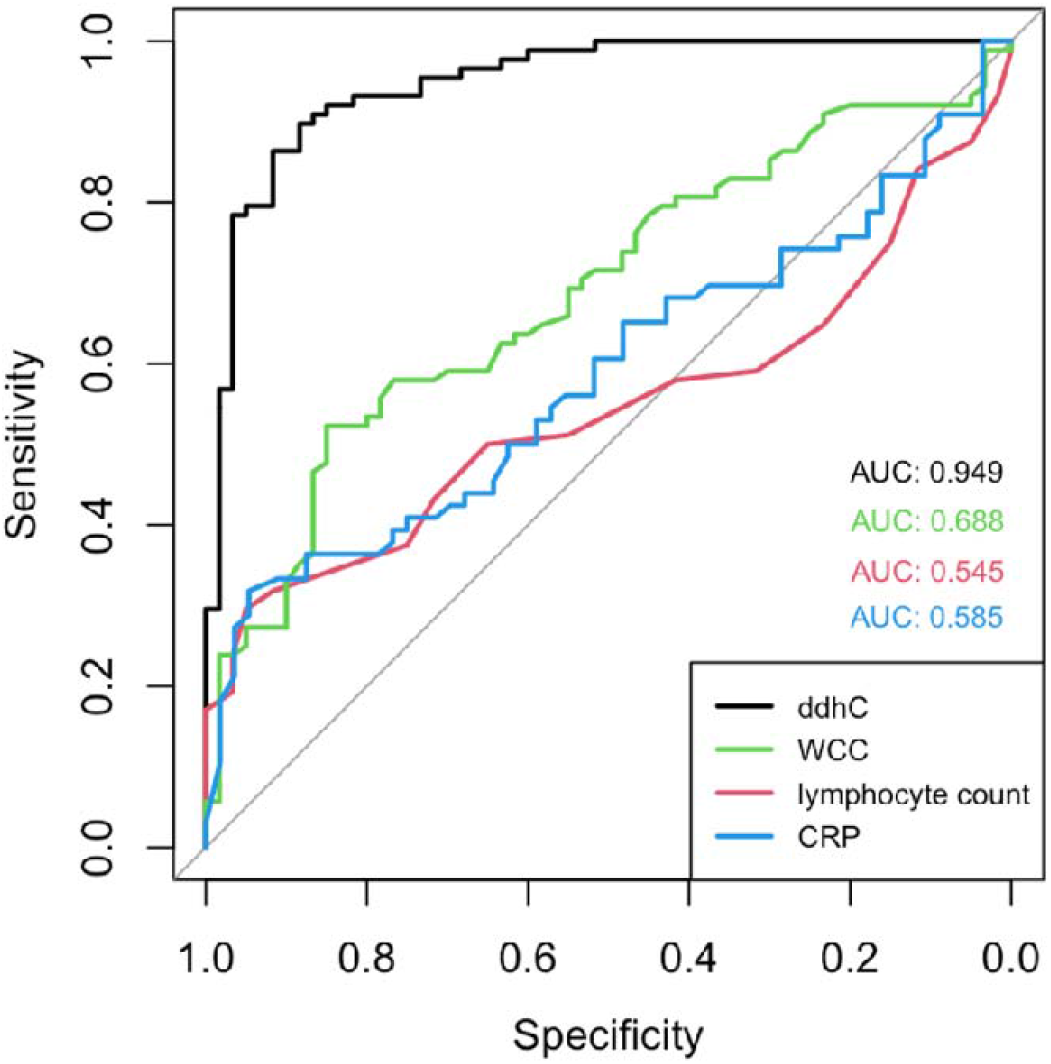
Comparison of area under the receiver operating characteristic curves (AUCs) between ddhC, white cell count (WCC), lymphocyte count and C-reactive protein (CRP) as biomarkers to distinguish viral infections from other groups in the primary analysis cohort. Black - ddhC (AUC = 0.949, n = 148); green - WCC (AUC = 0.688, n = 148); red - lymphocyte count (AUC = 0.545, n = 148); blue - CRP (AUC = 0.585, n = 122). Healthy controls not included as WCC, lymphocyte count and CRP not available.

### Interaction between ddhC and whole blood gene expression

RNA-Seq data was available from 122 patients in the HILIC+ primary analysis cohort (29 Gram-positive bacteraemia, 30 Gram-negative bacteraemia, 29 pre-COVID-19-viral, five COVID-19, 19 non-infected unwell control, 10 healthy controls). The correlation between log_2_-transformed ddhC intensity and counts for 18,248 genes was evaluated. The five genes with the highest correlation to ddhC intensity are shown in Supplementary Table 4, two of which are implicated in ddhCTP metabolism – *RSAD2* (viperin), aided by *CMPK2*, has been shown to mediate ddhCTP production during viral infection. ^41^ The correlation coefficient for viperin expression and ddhC intensity was 0.748 (p-value < 1×10^22^) and viperin was more highly expressed in patients with viral infections (Figures 6A and 6B). Data for *CMPK2* showed the same trends (Supplementary Figure 6).

**Figure 6.**
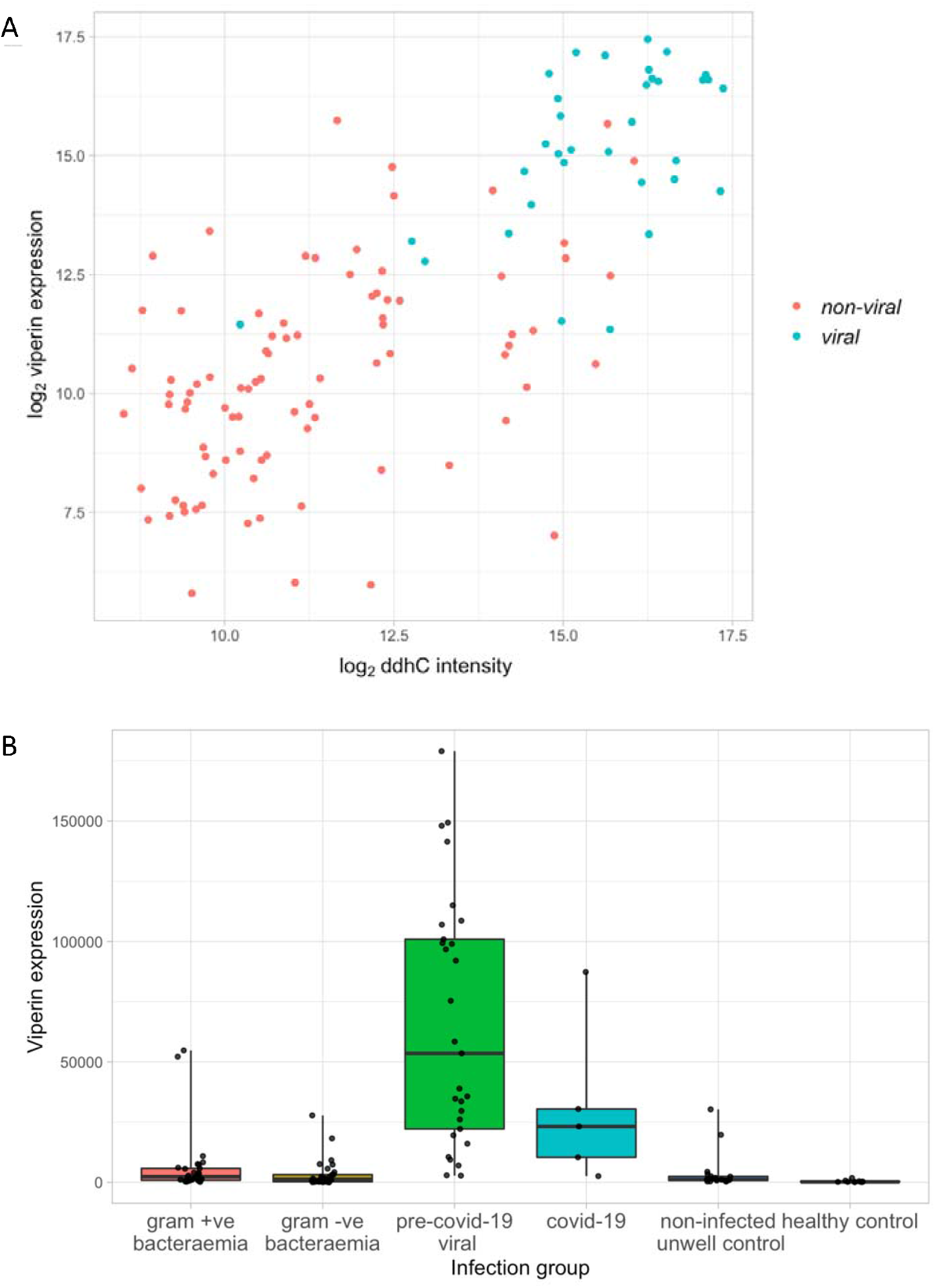
**A.** Correlation between ddhC intensity and viperin (*RSAD2*) gene expression in 122 patients. Non-viral = bacteraemic, non-infected unwell controls and healthy controls, viral = COVID-19 and pre-COVID-19 viral infection. Pearson correlation coefficient = 0.748, p-value < 1×10^22^. **B.** Viperin normalised gene counts for 122 patients in different infection groups.

## Discussion

The ability to rapidly differentiate infectious aetiologies is an urgent requirement, underlined by the ongoing COVID-19 pandemic. We capitalised on the sensitivity of high-resolution liquid chromatography coupled with mass spectrometry to discover that 3’-Deoxy-3’,4’-didehydro-cytidine (ddhC), a free base of the antiviral molecule ddhC-triphosphate (ddhCTP), was detectable in patient serum. ddhC was found to have a 36-fold higher median intensity in patients with viral infections, including COVID-19, compared to those with bacterial infections, non-infected inflammatory states and healthy controls, corresponding to an AUC of 0.954, sensitivity of 88.1% and specificity of 91.7%. It outperformed white cell count, lymphocyte count and CRP as a viral biomarker (AUCs of 0.688, 0.545 and 0.585, respectively).

ddhCTP has recently been shown to be the first and, to the best of our knowledge, only small molecule produced by humans that is capable of directly inhibiting viral replication machinery.^41^ Gizzi *et al*. showed that the enzyme viperin (virus inhibitory protein, endoplasmic reticulum-associated, interferon-inducible), aided by the genomically adjacent enzyme cytidylate monophosphate kinase 2 (*CMPK2*), catalyses the conversion of CTP to ddhCTP, which acts as a chain terminator for multiple viral RNA-dependent RNA polymerases (RdRPs). Synthetic ddhC traversed the plasma membrane of Vero and HEK293T cells, suggesting a mechanism for how ddhC might eventually reach the serum in detectable quantity. ddhC has also been detected in prokaryotic cells; *Escherichia coli* production of ddhCTP after viperin homolog expression was associated with T7 phage RdRP suppression, suggesting a role for ddhCTP in bacterial immunity to viruses.^23^ To our knowledge, ddhC has hitherto not been identified in humans or other mammals, nor associated with COVID-19. We showed that this antiviral molecule was a sensitive and specific serum biomarker for a range of viral infections, including COVID-19, in clinical samples of patients presenting to hospital. In a subset of patients for whom RNA-Seq data was available, we showed that viperin and *CMPK2* expression was also increased in patients with viral infections. Furthermore, of more than 18,000 genes, their expression was amongst the top five most highly correlated with ddhC intensity (correlation coefficients 0.75 and 0.76 respectively), providing a plausible mechanism by which ddhC may be produced during viral infection.

A robust serum biomarker of viral infection would provide real-time determination of infectious aetiology, aiding patient triage and decision-making regarding antimicrobial prescription. It would prove vital in infection prevention and control measures, especially in the context of a viral pandemic – where rapid detection of an acute viral illness, not dependent on nucleotide amplification via PCR, would enable prompt patient isolation. Further work is required to ascertain the role of ddhC as a biomarker in viral and bacterial co-infections as well as in chronic active viral infections.

The antiviral properties of ddhCTP rely on inhibition of viral RdRPs and are demonstrable *in vitro*, raising the possibility that it may have therapeutic action. RdRPs are an enticing target for novel antivirals and are an active focus of ongoing antiviral therapeutics research,^42^ as there are no functional homologues in uninfected human cells, thus off-target drug effects are less likely. Recently, Wood *et al*. demonstrated that ddhCTP can be robustly synthesised on a gram scale, facilitating further investigation of its use.^43^ Here we show that ddhC is produced naturally *in vivo* in response to viral infections, at levels that are detectable in the circulation, increasing the likelihood of an acceptable safety profile of ddhCTP as a therapeutic.

Our study demonstrates a number of strengths. We deliberately included both healthy and unhealthy non-infected controls, reducing the likelihood of selecting biomarkers confounded by inflammation unrelated to infection. We used stringent inclusion criteria for infected patients, excluding those where the timing of clinical presentation or PCR/culture result might have affected infection status at the point of sample acquisition. The study replicated ‘real-life’ sample collection, where serum samples may spend extended time outside of a fridge prior to biochemical analysis. We used admission-day samples taken prior to any intervention, the timepoint where a diagnostic test would be most useful.

Our study should be viewed in the context of its limitations. Firstly, in order to ensure diagnostic certainty, we only included the extremes of bacterial infection in the form of bacteraemia. Secondly, our cohort did not include fungal and protozoan infections. Thirdly, we only performed internal cross-validation, which will need further confirmation in an external cohort. We plan to address these limitations in future work to assess the performance of ddhC in new patient cohorts, including fungal and non-bacteraemic bacterial infections, as well as other infections seen outside of the UK. Fourthly, our cohort represents patients unwell enough to seek hospital attention – further work will be required to assess the role of ddhC in less unwell patients presenting to primary care and determine whether it is detectable in minimally invasive samples such as urine or saliva.

In conclusion, using high-fidelity metabolic profiling of serum from patients attending hospital, we found that the antiviral molecule ddhC is present in human serum during viral infection and represents an accurate biomarker for a wide range of viral infections, including COVID-19. If shown to perform consistently in further validation work, this biomarker will have a crucial role in the diagnostic repertoire for infectious diseases.

## Data Availability

All source data will be made publicly available through the European Bioinformatics Institute (EMBL-EBI) MetaboLights repository.

## Acknowledgments

The authors acknowledge the clinical research teams, diagnostic laboratory leads, and data managers whose work supports BioAID. The authors also acknowledge Ash Salam, Stephane Camuzeaux, Benjamin Cooper and Lynn Maslen, National Phenome Centre, for their technical and administrative expertise in running the LC-MS experiments, and Lachlan Coin for the development of the FS-PLS algorithm.

RM and TMR are NIHR Academic Clinical Fellows and with SS, and GSC, acknowledge support from the NIHR Imperial BRC, which also supports the Leonard and Dora Colebrook laboratory used for bioresourcing and the Imperial Tissue Bank; MK is a Sir Henry Wellcome Fellow (206508/Z/17/Z); HJ receives support from the Wellcome Trust (4-Year PhD programme, grant number 215214/Z/19/Z); HKL is a Medical Research Council (MRC) Clinical Research Training Fellow (MR/R502376/1); TMR acknowledges the Centre for Antimicrobial Optimisation, Imperial College London; GSC is an NIHR Research Professor; MN is a Wellcome Trust Investigator (207511/Z/17/Z) and acknowledges the NIHR UCLH BRC; SS acknowledges the NIHR Health Protection Research Unit (HPRU) in Healthcare-associated Infections & AMR; MRL, ZT, CS and EC receive support from the Medical Research Council (grant number MC_PC_12025) and metabolomics infrastructure support was provided by the NIHR Imperial BRC.

## Author contributions

SS, GSC and MN established the BioAID cohort. RM, SS, MRL, and ZT conceived and designed the study. RM, HKL and EM selected and prepared samples from the cohort for analysis. RM, PA, TMR, RH, MA, AH and DA acquired clinical data. CS facilitated LC-MS pre-processing. RM, HJ, MK and CS analysed data. EC facilitated metabolite identification. RM wrote the first draft of the manuscript. SS, MK, HJ, MRL, EC, MN, GC and CS edited the manuscript. All authors read and approved the final manuscript.

