## Supplementary Information for "Antiviral metabolite 3’-Deoxy-3’,4’-didehydro-cytidine is detectable in serum and identifies acute viral infections including COVID-19"

Figure 1A. Unsupervised unit-variance-scaled principal component analyses of the primary analysis cohort to assess for variance due to age and sex (n=161-163).

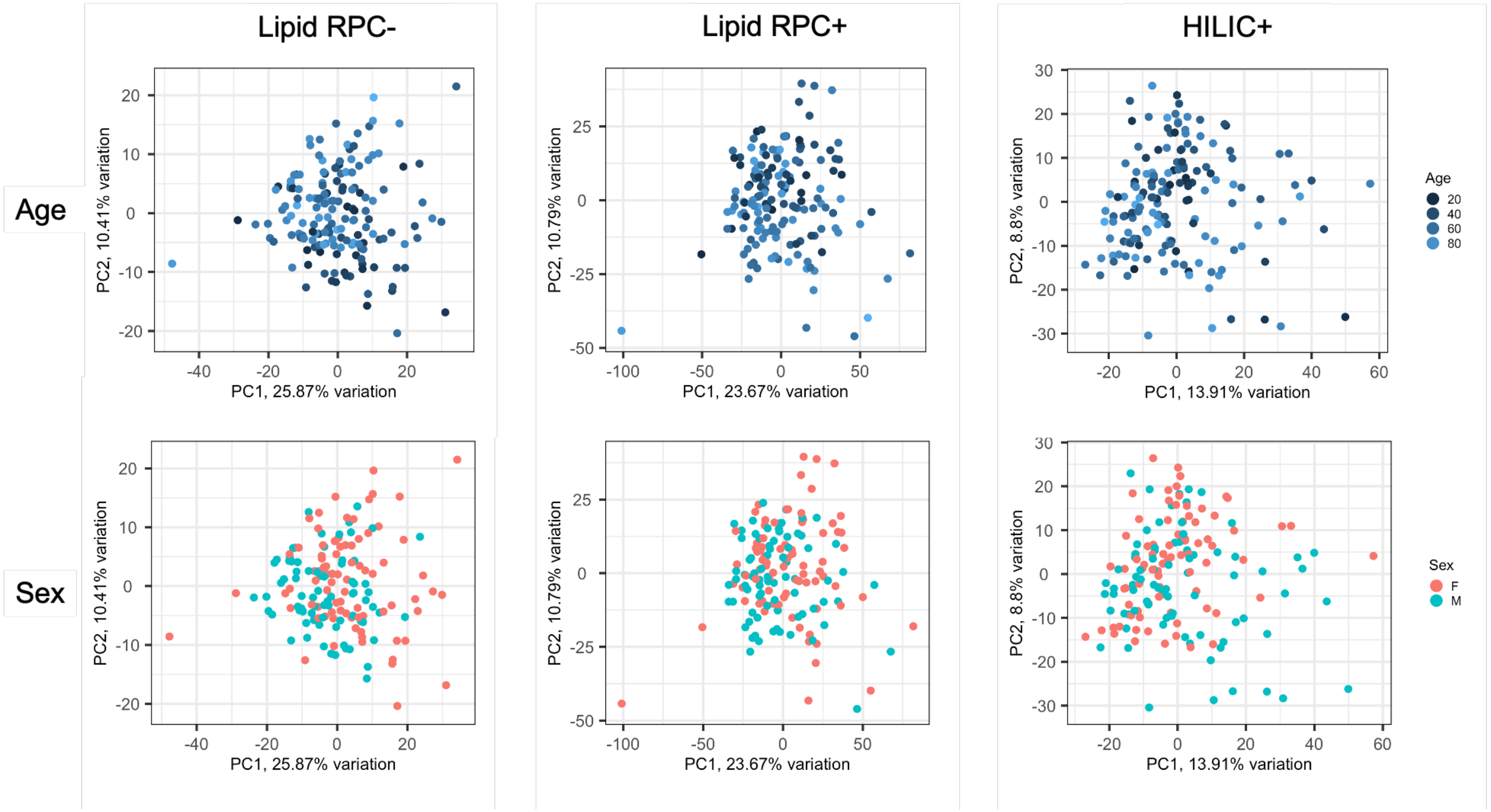

**Supplementary Figure 1A.** No variance in primary analysis cohort due to age or sex. HILIC = Hydrophilic interaction chromatography.

Figure 1B. Eigencor plots derived from principal component analyses of the primary analysis cohort to assess for variance due to age and sex (n=161-163).

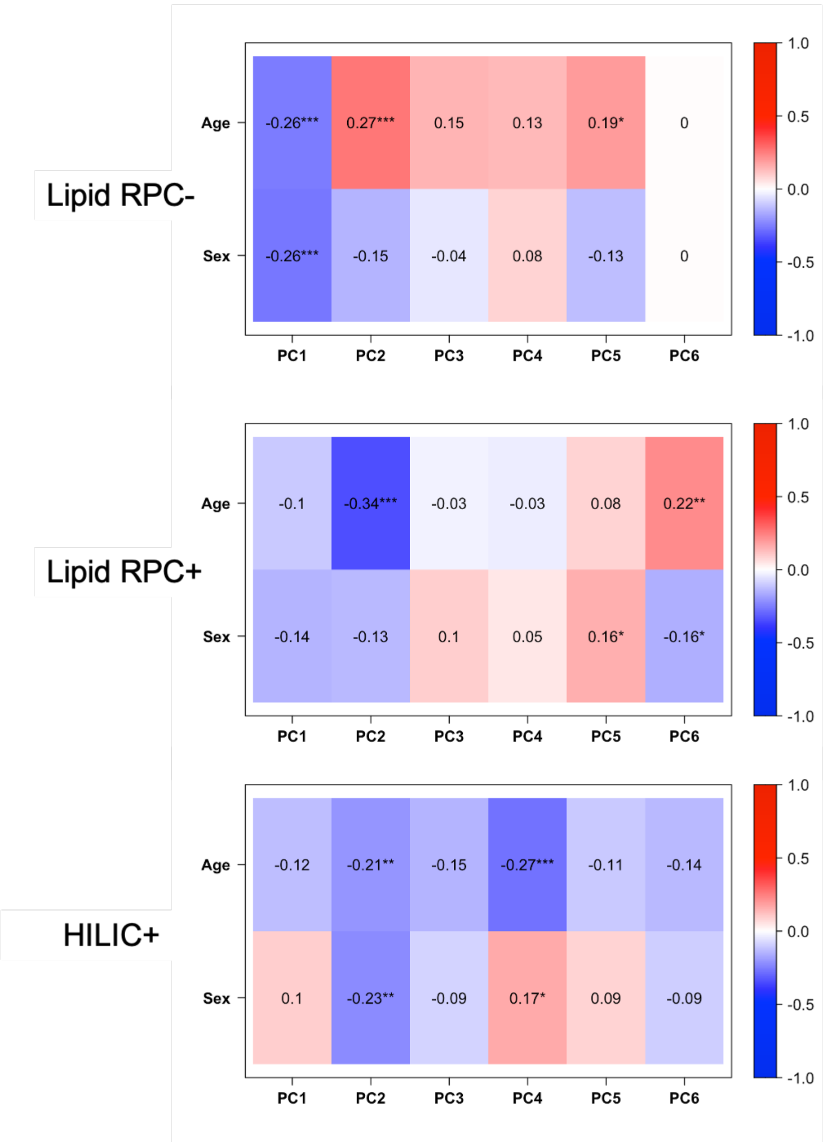

**Supplementary Figure 1B.** Minimal correlation between principal component variance and age or sex in primary analysis cohort. HILIC = Hydrophilic interaction chromatography. ***p-value<0.001, **<0.01, *<0.05.

### Figure 2. Univariate analyses for all comparisons in the primary analysis cohort (n=161-163).

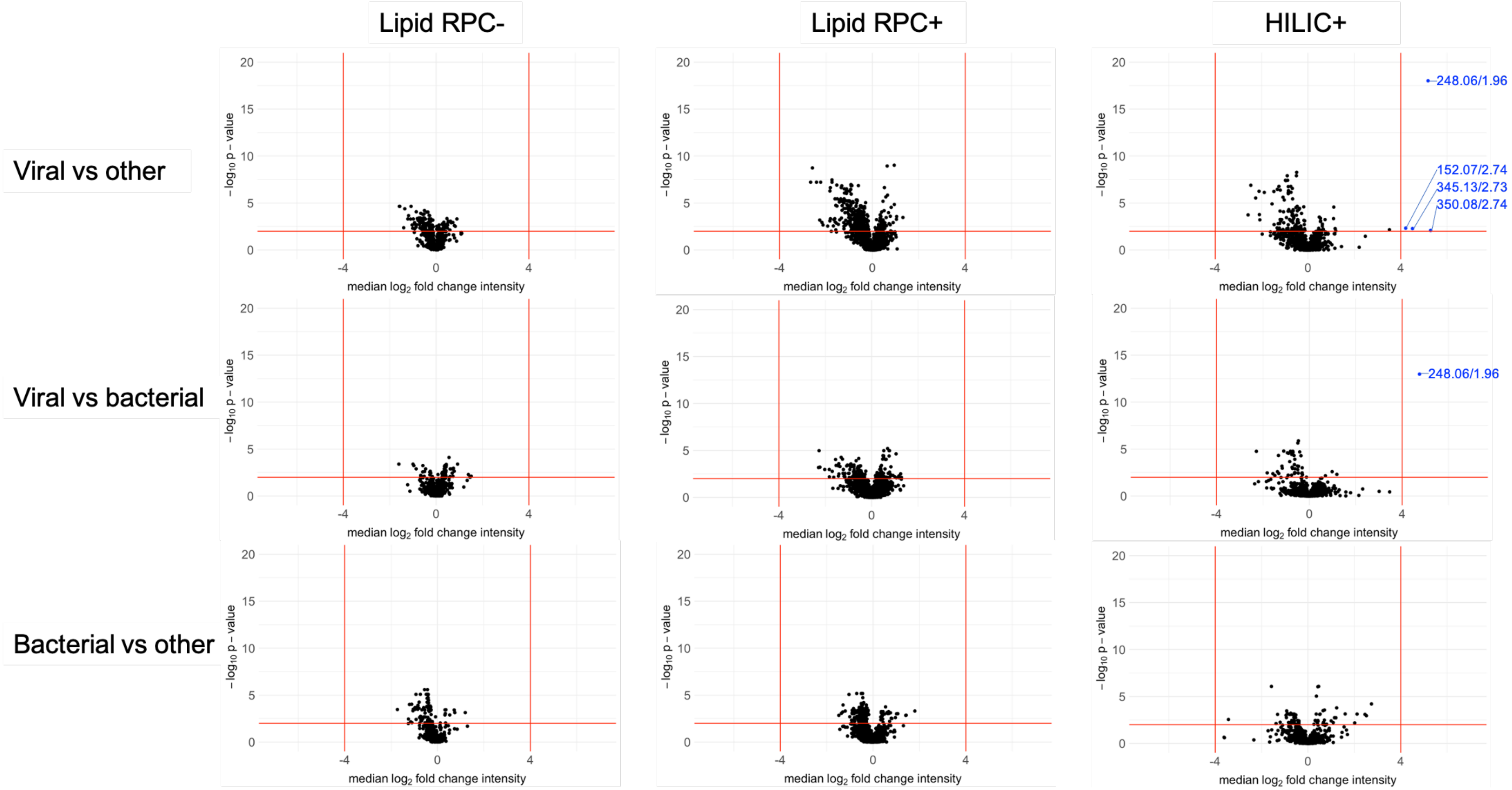

**Supplementary Figure 2.** Volcano plot showing median $\log_2$ fold change intensity of all features versus $-\log_{10}$ p-value in all assays when comparing: viral versus other, viral versus bacterial, and bacterial vs other. Threshold lines in red represent a fold-change of 16 [$\log_2$(foldchange) of 4] and p-value of 0.01 [$-\log_{10}$(p-value) of 2]. Candidate biomarkers are shown in blue by their mass:charge ratio/retention time.

### Figure 3. AUCs for ddhC in distinguishing viral versus other and viral versus bacterial groups; including samples excluded from the primary analysis due to prolonged time (>5 days) outside a -80°C freezer (n=239).

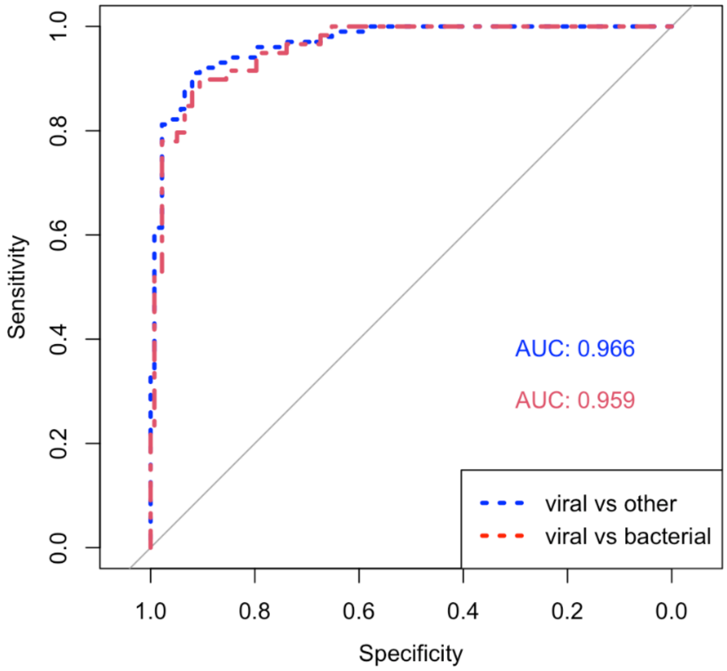

**Supplementary Figure 3.** AUCs for ddhC in distinguishing viral versus other and viral versus bacterial groups. Using all samples including those that spent >5days outside a -80°C freezer (n=239): **Blue.** AUC of 0.966 for ddhC (mass:charge ratio/retention time of 248.06/1.96) differentiating viral infections from all other groups. **Red.** AUC of 0.959 for ddhC differentiating viral from bacterial infection (controls omitted).

### Figure 4. Intensities for ddhC using dataset including samples excluded from the primary analysis due to prolonged time (>5 days) outside a -80°C freezer (n=239).

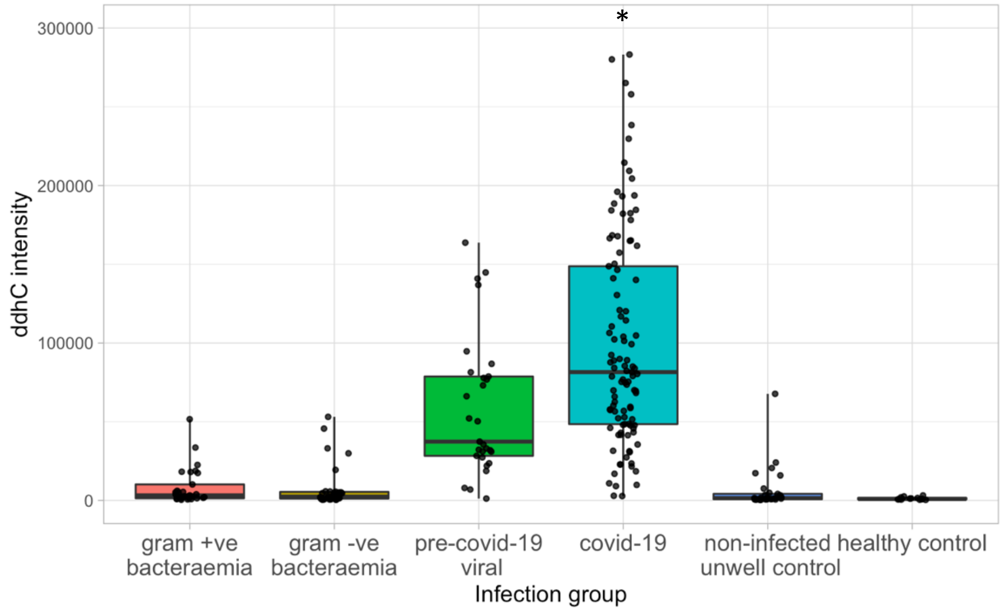

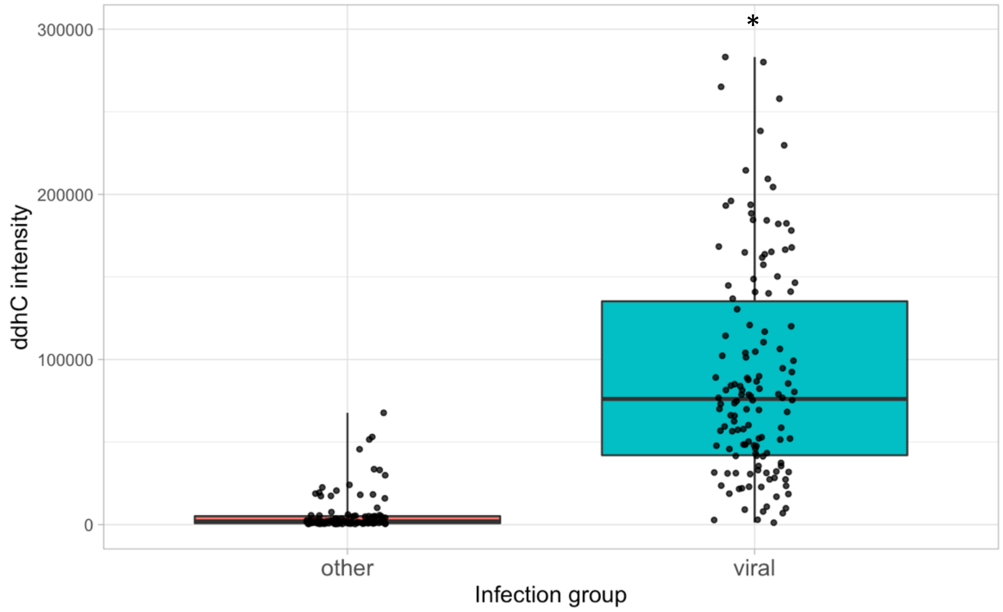

**Supplementary Figure 4.** ddhC (248.06/1.96) intensity data in different patient groups. Using all samples including those that spent >5days outside a -80°C freezer (n=239); points represent individual patients; boxes represent interquartile ranges with medians. **A.** All comparator groups. **B.** Pre-COVID-19 viral and COVID-19 grouped together into one 'viral' group vs all other groups. *4 samples in the COVID-19 group had an intensity of >400000, not shown.

### Figure 5. Effect of sex and age on ddhC intensity within viral group in the primary analysis cohort (n=60).

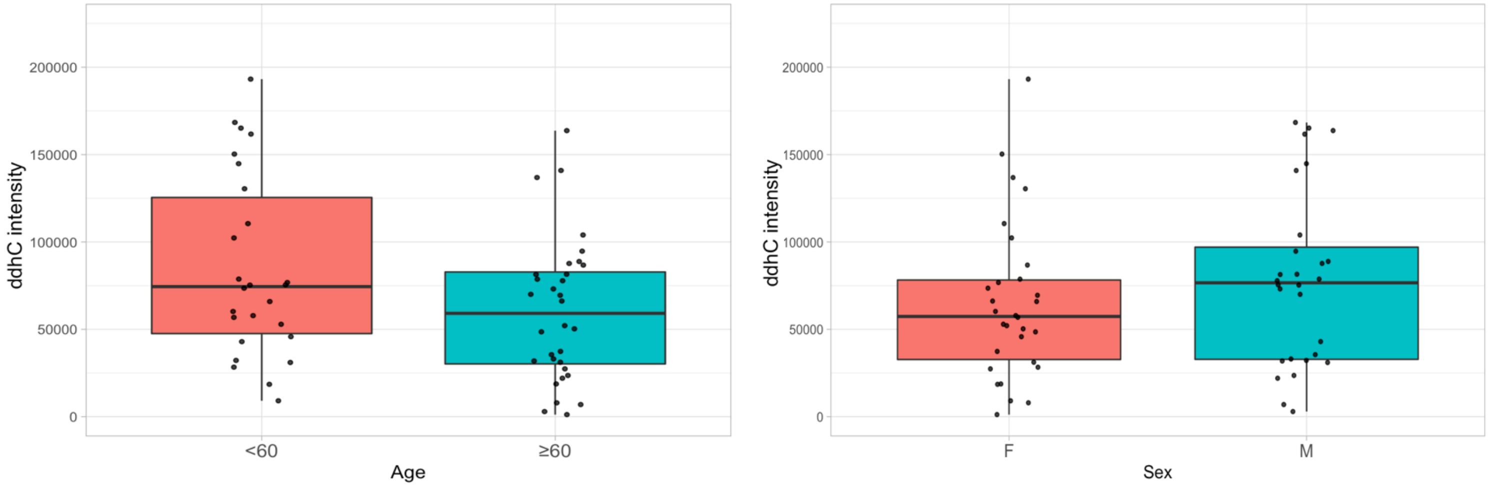

**Supplementary Figure 5.** No significant (p-value threshold <0.01) difference in ddhC intensity between age or sex within the viral (COVID-19 and pre-COVID-19 viral) group. 2 samples in the COVID-19 group had a ddhC intensity of >700000, not shown.

### Figure 6. Correlation between ddhC and *CMPK2* expression in different patient groups (n=122).

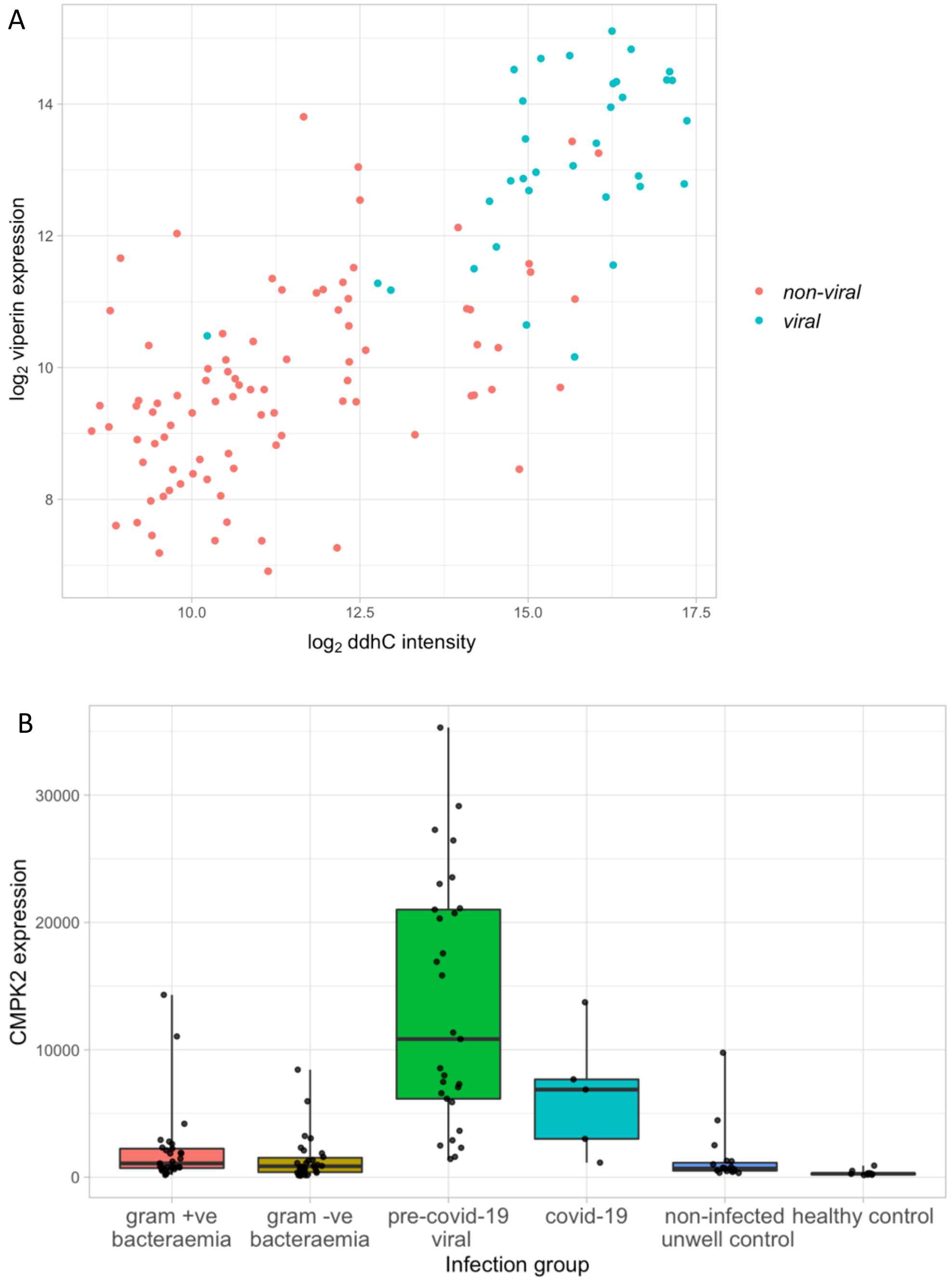

**Supplementary Figure 6. A.** Correlation between ddhC intensity and *CMPK2* gene counts in 122 patients. Non-viral = bacteraemic, non-infected unwell controls and healthy controls, viral = COVID-19 and pre-COVID-19 viral infection. Pearson correlation coefficient = 0.763, p-value < $1x10^{23}$. **B.** *CMPK2* expression in 122 patients in different infection categories.

### Table 1. Patient demographics and sample numbers for the primary analysis cohort.

| | Gram +ve bacteraemia | Gram -ve bacteraemia | Pre-COVID-19 viral | COVID-19 | Non-infected unwell control | Healthy control | Total |
|---|---|---|---|---|---|---|---|
| **n (included in at least one assay)** | 30 | 30 | 29 | 32 | 30 | 13 | 164 |
| **n (lipid RPC-)** | 30 | 30 | 29 | 31 | 30 | 13 | 163 |
| **n (lipid RPC+)** | 29 | 30 | 29 | 31 | 29 | 12 | 160 |
| **n (HILIC+)** | 29 | 30 | 29 | 31 | 29 | 13 | 161 |
| **Age (median [IQR])** | 71 [59-80] | 69 [60-79] | 41 [26-54] | 66 [60-80] | 52 [43-68] | 34 [31-45] | 60 [44-74] |
| **Female (n [%])** | 18 [60] | 14 [47] | 14 [48] | 18 [56] | 12 [40] | 8 [62] | 84 [51] |
| **WCC x10$^9$/L (median [IQR])** | 11.6 [7.3-14.8] | 13.5 [8.9-16.4] | 7.8 [5.1-9.5] | 7.2 [5.3-9.4] | 8.8 [6.6-12.2] | NA | 8.9 [6.2-12.9] |
| **Lymphocyte count x10$^9$/L (median [IQR])** | 0.9 [0.5-1.3] | 0.7 [0.5-1.2] | 0.6 [0.8-1.0] | 1.0 [0.9-1.3] | 1.8 [1.0-2.7] | NA | 0.9 [0.6-1.5] |
| **CRP mg/L (median [IQR])** | 56 [27-107] | 137 [42-205] | 31 [17-74] | 89 [50-154] | 6 [1-15] | NA | 56 [15-120] |

**Supplementary Table 1.** Sample numbers, demographics and routine biochemistry for the primary analysis cohort. IQR = interquartile range, HILIC = hydrophilic interaction chromatography, RPC = reversed-phase chromatography, WCC = white cell count, CRP = C-reactive protein. Routine biochemistry (WCC, lymphocyte count & CRP) not available for healthy controls, available for n = 151 (WCC & lymphocyte count) and n = 125 patients (CRP).

### Table 2. Confirmed infections in the primary analysis cohort.

| | Gram +ve bacteraemia | Gram -ve bacteraemia | Pre-COVID-19 viral | COVID-19 | Non-infected unwell control | Healthy control | Total |
|---|---|---|---|---|---|---|---|
| **n (total)** | 30 | 30 | 29 | 32 | 30 | 13 | 164 |
| ***Staphylococcus aureus*** | 10 | - | - | - | - | - | - |
| ***Streptococcus pneumoniae*** | 4 | - | - | - | - | - | - |
| ***B-haemolytic Streptococcus group C/G*** | 3 | - | - | - | - | - | - |
| ***Enterococcus faecalis*** | 3 | - | - | - | - | - | - |
| ***Streptococcus mitis*** | 2 | - | - | - | - | - | - |
| ***Streptococcus oralis*** | 1 | - | - | - | - | - | - |
| ***Streptococcus milleri*** | 1 | - | - | - | - | - | - |
| ***Streptococcus anginosus*** | 1 | - | - | - | - | - | - |
| ***B-haemolytic Streptococcus group B*** | 1 | - | - | - | - | - | - |
| ***Enterococcus casseliflavus*** | 1 | - | - | - | - | - | - |
| ***Listeria monocytogenes*** | 1 | - | - | - | - | - | - |
| ***Clostridium septicum*** | 1 | - | - | - | - | - | - |
| ***Lactobacillus paracasei*** | 1 | - | - | - | - | - | - |
| ***Escherichia coli*** | - | 19 | - | - | - | - | - |
| ***Klebsiella pneumoniae*** | - | 3 | - | - | - | - | - |
| ***Proteus mirabilis*** | - | 2 | - | - | - | - | - |
| ***Enterobacter cloacae*** | - | 2 | - | - | - | - | - |
| ***Klebsiella oxytoca*** | - | 1 | - | - | - | - | - |
| ***Serratia marcescens*** | - | 1 | - | - | - | - | - |
| ***Moraxella catarrhalis*** | - | 1 | - | - | - | - | - |
| ***Haemophilus parainfluenzae*** | - | 1 | - | - | - | - | - |
| **Influenza A** | - | - | 11 | - | - | - | - |
| **Adenovirus** | - | - | 5 | - | - | - | - |
| **Measles** | - | - | 3 | - | - | - | - |
| **Varicella Zoster Virus** | - | - | 3 | - | - | - | - |
| **Dengue** | - | - | 2 | - | - | - | - |
| **Herpes Simplex Virus** | - | - | 1 | - | - | - | - |
| **Rhinovirus / Adenovirus / Parainf 1*** | - | - | 1 | - | - | - | - |
| **Parainfluenza 2** | - | - | 1 | - | - | - | - |
| **Herpes Simplex Virus + Adenovirus** | - | - | 1 | - | - | - | - |
| **Enterovirus** | - | - | 1 | - | - | - | - |
| **SARS-CoV-2** | - | - | - | 32 | - | - | - |

**Supplementary Table 2.** Confirmed infections for the primary analysis cohort. *All three of rhinovirus, parainfluenza 1 virus and adenovirus (weak positive) were detected on a respiratory viral PCR assay for this patient.

### Table 3. Cross-validation of feature performance using FS-PLS.

| | Assay | Most frequent feature | FS-PLS runs (/100) | Median test AUC | IQR test AUC |
|---|---|---|---|---|---|
| **Viral vs other** | Lipid RPC+ | 778.54/5.01 | 61 | 0.843 | 0.809-0.867 |
| | Lipid RPC- | 766.53/5.76 | 46 | 0.775 | 0.724-0.802 |
| | HILIC+ | 248.06/1.96 | 100 | 0.957 | 0.943-0.970 |
| **Viral vs bacterial** | Lipid RPC+ | 711.58/5.90 | 37 | 0.833 | 0.796-0.867 |
| | Lipid RPC- | 717.53/6.00 | 39 | 0.731 | 0.701-0.779 |
| | HILIC+ | 248.06/1.96 | 99 | 0.951 | 0.926-0.969 |
| **Bacterial vs other** | Lipid RPC+ | 740.61/6.62 | 31 | 0.741 | 0.710-0.794 |
| | Lipid RPC- | 871.69/8.09 | 30 | 0.768 | 0.726-0.813 |
| | HILIC+ | 512.34/5.09 | 69 | 0.797 | 0.767-0.839 |

**Supplementary Table 3.** Cross-validation of feature performance in differentiating viral vs other, bacteraemic vs viral and bacteraemic vs other patient groups using 100 forward selection-partial least squares (FS-PLS) runs comprising 70:30 training:test splits. FS-PLS runs (/100) = the number of times the feature was selected in 100 FS-PLS runs. ddhC (248.06/1.96) was selected as the discriminating feature in all 100 FS-PLS runs in the HILIC+ assay comparing viral vs other, generating a median test area under the receiver operating characteristic curve (AUC) of 0.957. Features are shown by mass:charge ratio/retention time. IQR = interquartile range, RPC = reversed-phase chromatography, HILIC = hydrophilic interaction chromatography.

### Table 4. Five genes most highly correlated with ddhC.

| Gene | Ensembl ID | Correlation with ddhC | p-value |
|---|---|---|---|
| ***CMPK2*** | ENSG00000134326 | 0.763 | $< 1x10^{23}$ |
| ***SPATS2L*** | ENSG00000196141 | 0.759 | $< 1x10^{23}$ |
| ***IFI27*** | ENSG00000165949 | 0.756 | $< 1x10^{23}$ |
| ***RSAD2 (viperin)*** | ENSG00000134321 | 0.748 | $< 1x10^{22}$ |
| ***Unnamed**** | ENSG00000233785 | 0.743 | $< 1x10^{21}$ |

**Supplementary Table 4.** Pearson's correlation coefficients between $log_2$-transformed whole blood-derived gene expression and $log_2$-transformed ddhC intensity, with the five (of 18,248) most highly correlated genes shown. *No identification available.

### Metabolite identification

For the feature identified as a discriminator for viral infections (mass:charge ratio (*m/z*) of 248.0647 and retention time of 1.96 mins), we used an in-house R-script to assign ion type $[M+Na]^+$. We extracted intensity values for four other features representing the same metabolite (eluting at the same retention time and having identical chromatographic peak shapes), namely the first isotope of $[M+Na]^+$ ion at *m/z* 249.0676 , $[M+H]^+$ at *m/z* 226.0827, $[M+K]^+$ at *m/z* 264.0383, and in-source fragment at *m/z* 112.0517. All correlated features gave comparable individual AUCs to the identified feature (AUC 0.954) when comparing viral versus all other groups.

| Ion type | Mass:charge ratio | AUC viral vs all other groups |
|---|---|---|
| **First isotope $[M+Na]^+$** | 249.0676 | 0.943 |
| **$[M+H]^+$** | 226.0827 | 0.948 |
| **$[M+K]^+$** | 264.0383 | 0.951 |
| **In-source fragment** | 112.0517 | 0.953 |

**Supplementary Table 2.** AUCs when comparing viral versus all other groups in the HILIC+ data set for the primary analysis cohort (n = 161) for all features representing the same metabolite as the identified feature of interest (248.06/1.96).

The molecular formula of the metabolite of interest was determined to be C9H11N3O4 through matching the mass of the $[M+H]^+$ ion to chemical and spectral databases. We performed MS/MS analysis of the parent $[M+H]^+$ ion and $[M+Na]^+$.

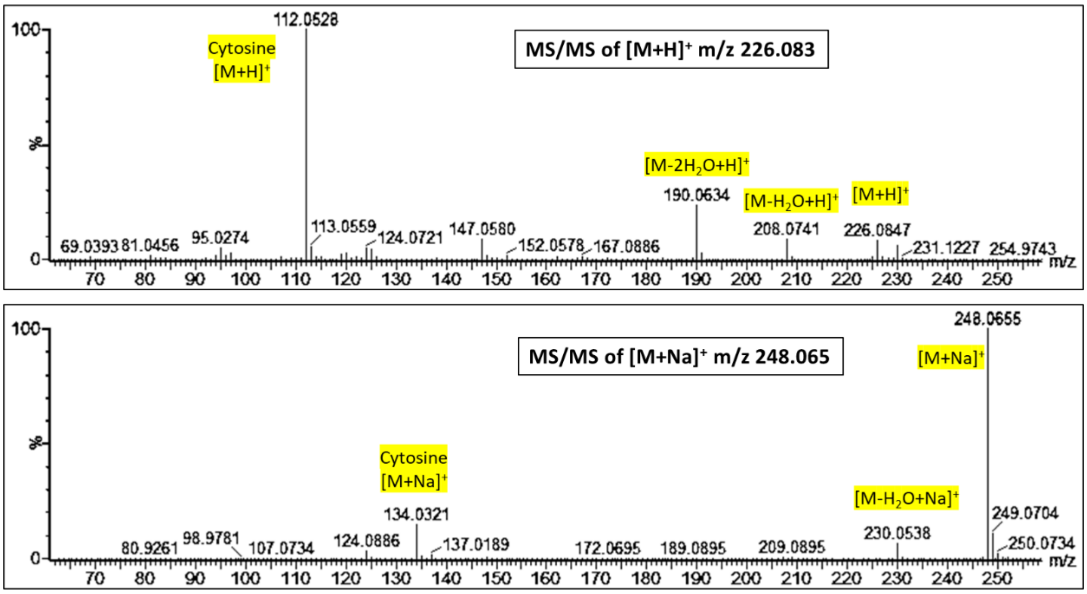

**Supplementary Figure 7.** MS/MS spectrum obtained with collision energy ramp of 10-45V for $[M+H]^+$ ion *m/z* 226.0827 and $[M+Na]^+$ ion *m/z* 248.0647 using a study sample with high intensity of these features.

The experimental MS/MS spectrum for the $[M+H]^+$ ion species (*m/z* 226.0827) revealed the loss of one and two water moieties (*m/z* 208.072 and *m/z* 190.062, respectively) and the fragment with *m/z* 112.05 consistent with cytosine ($[M+H]^+$). The experimental MS/MS spectrum for the $[M+Na]^+$ ion species (*m/z* 248.0647) revealed analogous species.

No matches were found for these spectra in databases (Human Metabolome Database, METLIN, NIST17, Mass Bank of North America). We therefore hypothesized that the metabolite of interest was a nucleoside previously reported in the literature – 3'-Deoxy-3',4'-didehydro-cytidine (ddhC), a free base of the antiviral ribonucleotide 3'-Deoxy-3',4'-didehydro-cytidine triphosphate.[1] The experimental MS/MS spectrum of $[M+H]^+$ obtained in our study samples matched the previously published fragmentation pattern of natural ddhC detected in cell lysates of *E. coli* and authentic chemical standard of ddhC.[2]

Finally, the structure of metabolite was definitively identified by analysing a chemical standard of ddhC (acquired from Berry & Associates) in parallel with the study samples, using the HILIC profiling method and tandem MS/MS to match chromatographic retention time and fragmentation spectrum, respectively. For the former, a spike-in technique was used, where the pooled serum sample was mixed with different concentrations of the ddhC chemical standard (2.5, 5 and 10 ng/mL).

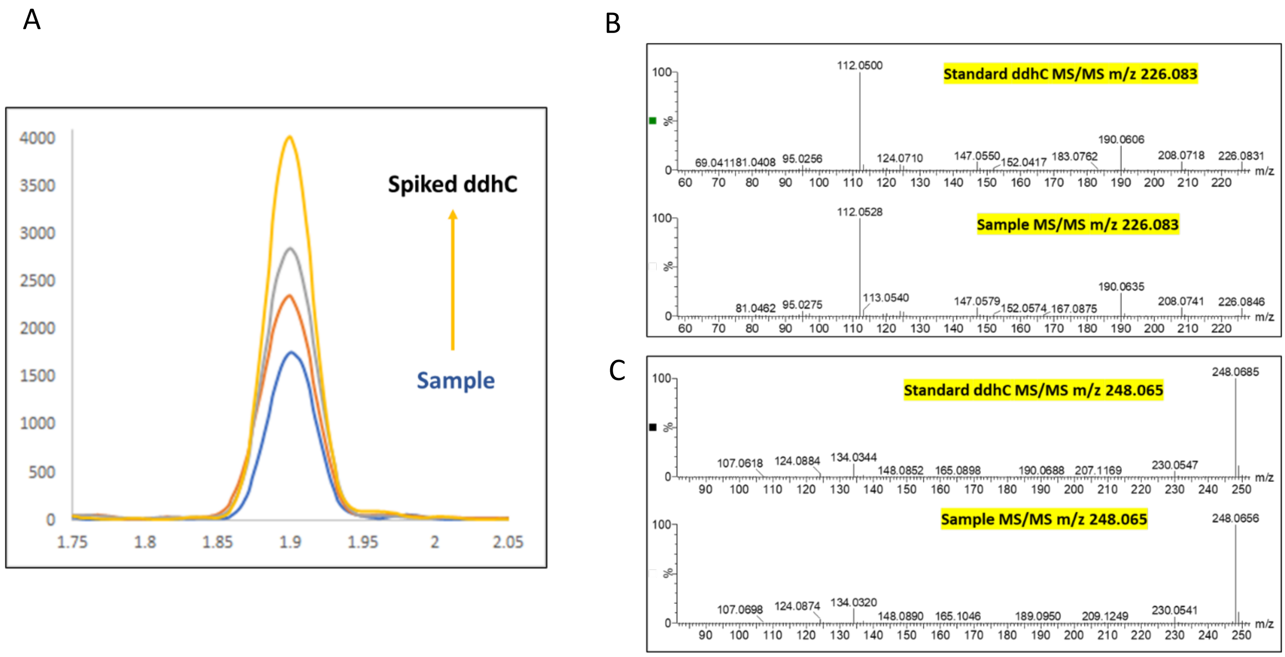

**Supplementary Figure 8.** Definitive identification of ddhC using a chemical standard. **A.** HILIC-derived chromatographic retention time of the feature of interest in the pooled sample (blue) matches that of the pooled serum sample spiked with different concentrations of ddhC chemical standard (red = 2.5ng/mL, grey = 5ng/mL, yellow = 10ng/mL). **B & C.** MS/MS spectra of sample $[M+H]^+$ and $[M+Na]^+$ ions match the fragmentation patterns of the ddhC standard.
